# Lead exposure and preeclampsia in sub-Saharan Africa: modelling the potential contribution to maternal disease burden and opportunities for prevention

**DOI:** 10.64898/2026.01.02.25343170

**Authors:** Mark A. S. Laidlaw

**Affiliations:** Independent Researcher.

**Author notes:** Corresponding author: Mark A. S. Laidlaw.

## Abstract

**Background:** Hypertensive disorders of pregnancy, particularly preeclampsia, are a leading cause of maternal morbidity and mortality in sub-Saharan Africa (SSA). Although environmental lead exposure remains widespread across the region, its contribution to the burden of preeclampsia has not previously been quantified at the population level.

**Methods:** We conducted a modelling study using maternal blood lead level (BLL) data synthesised from pregnancy biomonitoring studies in SSA and supplemented these with soil-derived exposure scenarios representing severely contaminated environments. Biomonitoring data represented real-world exposure distributions, whereas soil-derived scenarios evaluated potential impacts in highly contaminated communities. Published meta-analytic evidence indicating a 1.6% increase in the odds of preeclampsia per 1 µg/dL increase in maternal BLL was used to estimate predicted preeclampsia risk across plausible baseline prevalence scenarios. Sensitivity analyses evaluated uncertainty associated with exposure extrapolation and baseline prevalence, and population attributable fractions (PAFs) were estimated.

**Results:** Maternal BLLs reported in SSA pregnancy studies ranged from 0.83 to 99.0 µg/dL, with a study-level mean of 26.24 µg/dL, approximately 10–40 times higher than maternal BLLs reported in North America, Europe and other high-income regions. Biomonitoring-derived BLLs were associated with modest but consistent increases in predicted preeclampsia risk. Estimated PAFs suggested that, if the observed association is causal, lead exposure could account for approximately 6–8% of preeclampsia cases at low exposure (∼5 µg/dL), 19–21% at typical urban exposure (∼15 µg/dL), and 41–47% in highly exposed populations (∼40 µg/dL). Relative exposure-response relationships remained robust across sensitivity analyses.

**Conclusions:** Maternal lead exposure is a potentially modifiable environmental risk factor for preeclampsia in SSA that has received little attention in maternal health policy. These findings support integrating lead exposure prevention, environmental remediation, and targeted maternal biomonitoring into maternal health programmes, particularly in high-risk settings, to reduce preventable preeclampsia and improve maternal health outcomes.

## Introduction

Preeclampsia is a leading cause of maternal morbidity and mortality worldwide with a disproportionate burden in low and middle-income countries, particularly sub-Saharan Africa (SSA) and Southern Asia (WHO, 2025).

Preeclampsia is a pregnancy complication usually occurring after 20 weeks’ gestation and is defined by new-onset high blood pressure with signs of organ involvement. Symptoms may range from mild to severe and can develop suddenly. Many people may have no noticeable symptoms early on, which is why regular antenatal checks are essential. Severe preeclampsia can also affect blood-clotting, liver, kidney, and brain function, and may progress without obvious early symptoms.(WHO, 2025). Some of the risk factors include pre-existing conditions such as hypertension, diabetes, or kidney disease, first time pregnancies, obesity, multiple pregnancies (twins, triplets, etc.) and family history of pre-eclampsia (WHO, 2025).

Common signs of preeclampsia include high blood pressure, proteinuria, sudden swelling of the face, hands and feet, severe headache, visual disturbance, upper abdominal or right-upper-quadrant paint, nausea and vomiting and feeling unwell or short of breath (WHO, 2025).

Eclampsia is the most severe form of preeclampsia and is diagnosed when a person with preeclampsia develops seizures that cannot be explained by other causes. Eclampsia is a medical emergency due to the risk of maternal and fetal injury or death, requiring urgent hospital care.

The pooled incidence of preeclampsia in South Saharan Africa (SSA) is approximately 13% (95% CI 0.12–0.14) across included studies (Jikamo et al, 2023). The World Health Organization (WHO, 2025) estimated that in 2023, there were approximately 182,000 maternal deaths in SSA, or 70% of the total 260,000 of women’s deaths during and following pregnancy. It has been estimated that approximately 16% of these deaths were due to hypertensive disorders of pregnancy (HDP). This implies that there were approximately 29,000 HDP-related maternal deaths annually in the SSA (with pre-eclampsia/eclampsia constituting a substantial fraction).

Established maternal, clinical, nutritional, socioeconomic and health-system factors for preeclampsia include maternal age, parity, obesity, diabetes, hypertension and access to antenatal care (Duckett and Harrington, 2005; Abolos et al., 2013).

### Lead Exposure

The main sources of lead exposure globally are mining and smelting, lead-acid battery manufacture and recycling, lead-containing paints and consumer products, contaminated food, soil and dust, and drinking water contaminated by lead plumbing (WHO, 2026). Lead in soil and soil dust are typically derived from past petrol lead emissions (Dietrich et al., 2026), deteriorating exterior lead based paints (Laidlaw et al., 2018)., industry, smelters (Ettler, 2016), used lead acid battery (ULAB) recycling (Ericson et al., 2016) and fill material contaminated with lead from various sources.

In most high-income nations, lead began to be used in petrol beginning in 1920’s, peaking in the 1970’s and a ban took effect throughout the 1980s. However, this polluting fuel was still in use across virtually all low-and middle-income countries, along with a few OECD members until 2002. The UN Environment Program (UNEP) initiated efforts to achieve the global elimination of lead in petrol in 2002, and lead in petrol was eliminated globally in 2021 (UNEP, 2021;). During the time that lead was being emitted from petrol emissions, blood lead levels of entire populations closely followed atmospheric lead levels (Kristensen, 2015; Angrand et al., 2022), indicating that the main source of lead exposure during this time was from lead in petrol.

In the United States, Mielke et al. (2011) reported that between 1950 and 1982 approximately 4.6 million tons of lead in petrol was emitted into urban areas in the USA (Mielke et al, 2011). The amount of lead emitted into the atmosphere in SSA has not been publicly reported, however the lead content in gasoline in Africa has been reported as approximately 1.9–3.0 grams of lead per US gallon (0.5–0.8 g lead per litre (g Pb/L)) (Maresky and Grobler, 1993. Thomas and Kwong (2001) estimated that about 9000 tonnes per year of lead was used in gasoline in Africa, with more than half used in South Africa and Nigeria.

Lead in petrol was emitted into the atmosphere of urban areas where it accumulated in surface soils (Mielke and Reagan, 1998) in areas with high historical traffic volumes (Dietrich et al., 2026), and in soils adjacent to older homes that used exterior lead-based paint (Laidlaw et al., 2018) as well as adjacent to homes with brick exteriors from lead in petrol (Mielke et al., 1983). In inner city areas, soil lead concentrations are typically highest and decay with distance away from roadways (Laidlaw and Filippelli, 2008; Dietrich et al., 2026) and house side (Mielke et al., 1983). Soil lead levels typically formed “bullseye” patterns in urban areas, with highest concentrations in the inner cities with concentrations decaying with distance away from the inner-city areas. This has been demonstrated in American cities such as New Orleans (Mielke et al, 2019), Chicago (Thorstenson et al., 2025), Indianapolis (Filippelli et al, 2005), and London in the UK (BGS, 2020). This same pattern of soil lead contamination is expected in large cities of SSA and warrants further assessment.

Lead in soil dust, the atmosphere and children’s blood levels vary seasonally with peaks in the dry periods due to soil resuspension (Zahran et al., 2013; Laidlaw et al., 2014). In north America, this typically occurs in the summer and autumn, however in Mexico lead in soil is resuspended in the winter when soils are dryest (Soto-Jiménez et al., 2026). Heavily lead contaminated roadside soil dust has also been shown to resuspended into the atmosphere due to atmospheric turbulence from passing vehicles as was shown in London, England (Resongles et al., 2021). It is likely that these same processes are occurring in SSA.

### General Lead Exposure in South Saharan Africa

Substantial lead exposure persists across sub-Saharan Africa due to petrol legacy soil and dust contamination, soil contamination from deteriorating exterior lead based paints, dust from deterioration of interior lead-base paints, mining and smelting activities, informal recycling of lead–acid batteries, contaminated consumer products and food and drinking water (UNICEF and Pure Earth, 2020). Major sources of lead exposure in SSA are presented in **Table 1**, below.

**Table 1.** Ranked main sources of lead exposure in SSA (adapted from UNICEF and Pure Earth, 2020).

| Rank | Source category | Typical exposure intensity | Population most affected |
| --- | --- | --- | --- |
| 1 | Artisanal & informal mining | Very high | Women, children, entire communities |
| 2 | Informal battery recycling & smelting | Very high | Whole households, workers' families |
| 3 | Leaded paints (household & industrial) | High | Children, pregnant women |
| 4 | Contaminated consumer products & traditional items | High (variable) | Women, infants |
| 5 | Legacy leaded-petrol soil & urban dust | Moderate-high | Urban residents, children |
| 6 | Occupational exposure in informal industries | Moderate-high | Working women |
| 7 | Food and drinking water contamination | Moderate - high | General population |

### Lead Levels in Soil of SSA

Studies across sub-Saharan Africa have documented substantial spatial variation in soil lead concentrations, with elevated levels in urban residential and roadside environments and particularly severe contamination associated with mining, ore processing and battery recycling. The studies summarised in **Table 2** demonstrate the potential for substantial contamination of environments where people live and work; however, differences in site selection, sampling and analytical methods limit direct comparisons, and findings from selected contaminated locations should not be interpreted as representative national or regional concentrations.

**Table 2.** Selected studies of soil lead concentrations in sub-Saharan Africa. Concentrations are expressed as mg/kg unless an exceedance statistic is reported. These selected studies describe residential, roadside and source-affected settings and do not represent national or regional average concentrations.

| Location and study | Sampling setting | Reported soil lead concentrations | Main finding and interpretation |
| --- | --- | --- | --- |
| **Kabwe, Zambia**<br>**Residential study published in Sakala (2026)** | <u>Homes of pregnant women</u> in four townships; 510 households | Median 389.8; IQR 53.4–2,044.6; 48.8% exceeded the study comparison value of 400 | Relevant to maternal exposure research. Soil concentrations alone do not establish maternal absorbed dose. |
| **Johannesburg, South Africa**<br>**Mathee et al. (2018)** | Residential gardens in four neighbourhoods | Inner-city suburb: mean 241.7; median 158.1; range 0.1–2,141 | Substantial residential contamination. These statistics describe the inner-city suburb, not all four neighbourhoods combined. |
| **Addis Ababa, Ethiopia**<br>**Teju et al. (2012)** | 45 roadside soil samples from 14 sites and a remote control sample | Roadside average $418.6 \pm 3.4$; control $18.8 \pm 0.5$ | Authors considered historical leaded petrol an important explanation for roadside enrichment; petrol was not established as the sole source. |
| **Dareta, Zamfara, Nigeria**<br>**Udiba et al. (2019)** | Village affected by lead-rich gold-ore processing; sampled June–July 2013 after earlier remediation | Means: residential compounds 1,029.4; market square 1,524.0; old grinding mills 1,404.6; new grinding mills 6,724.7 | Severe contamination remained in residential and processing areas, demonstrating the need to control continuing sources after cleanup. |
| **Thiaroye-sur-Mer, Dakar, Senegal**<br>**Haefliger et al. (2009)** | Community involved in informal used lead-acid battery recycling | Outdoor soil maximum 302,000 | Extreme hotspot associated with severe community lead poisoning. The maximum is not representative of typical Dakar soil. |
| **Multiple African countries**<br>**Gottesfeld et al. (2018)** | 118 samples at 15 battery-recycling plants and one manufacturing facility | Overall range <40–140,000; arithmetic mean in surrounding communities approximately 2,600 | Includes Cameroon, Ghana, Kenya, Mozambique, Nigeria and Tanzania, plus Tunisia. Pooled results are therefore not exclusively sub-Saharan estimates. |

Notes: mg/kg and µg/g are numerically equivalent. IQR = interquartile range. Means, medians, maxima and exceedance proportions are distinct statistics and should not be pooled as equivalent observations. Study comparison values are not universal safety thresholds.

### Lead Levels in Food in SSA

Using 25 total diet studies covering 46 countries and biokinetic modelling, Kinally et al. (2026) estimated that dietary lead may contribute approximately 40–50% of global lead exposure, supporting its role as a major exposure source. Studies in sub-Saharan Africa have documented elevated lead concentrations in vegetables, staple crops and prepared foods, although contamination varies considerably between locations and food types. Reported findings include a mean vegetable lead concentration of 2.78 mg/kg in Lagos, Nigeria, mean concentrations of 3.30–11.25 mg/kg in cooked street-vended foods in Kumasi, Ghana, and concentrations of 27.3–29.0 mg/kg dry weight in maize grain grown on unamended, mining-contaminated soil near Kabwe, Zambia (Ibukun et al., 2010; Ankar-Brewoo et al., 2020; Mwilola et al., 2020). Evidence identifies atmospheric deposition onto edible leaves and contamination from artisanal cookware as additional pathways beyond root uptake (Nabulo et al., 2006; Jitaru et al., 2019). Lead-contaminated spices, particularly turmeric adulterated with lead chromate pigments, can also constitute a substantial source of human lead exposure in affected populations (Forsyth et al., 2023).”

### Lead Levels in Water in SSA

The data regarding lead levels in water of SSA is sparse and no studies regarding the association between eclampsia or preeclampsia and lead in water in SSA have been located. Across SSA, reported lead concentrations in groundwater and borehole-derived drinking water vary widely, from <5 µg/L in relatively uncontaminated rural aquifers to >100 µg/L in mining-impacted regions such as Kabwe, Zambia (Siame et al., 2024). Multi-country evidence from West Africa indicates that exceedances of the WHO guideline (10 µg/L) occur in approximately 9% of drinking-water systems, with contamination frequently attributable to brass plumbing components rather than aquifer geochemistry (Fisher et al., 2021). In contrast, studies in Nigeria (Tyopine et al., 2024) and Zambia (Siame et al., 2024) demonstrate substantially higher concentrations (mean ∼20 µg/L and median >100 µg/L, respectively) in areas affected by lead–zinc mining, where exceedances are widespread or universal. Overall, the literature highlights a critical distinction between infrastructure-derived lead in borehole systems and true groundwater contamination, with the latter primarily associated with mining and waste sources.

### Relationship Between Soil Lead and Blood Lead

Soil lead levels have been shown to be correlated with children’s blood lead levels. In New Orleans, Louisiana (USA), Mielke et al. (2007)used a dataset of 55,551 children and a soil lead dataset of 5,467 children and found that the relationship between soil lead (SL)and children’s pooled blood lead level was curvilinear (r^2^ of 0.528; P-value of 1.0 × 10^− 211^). Below 100 mg/kg SL children’s BL exposure response was steep (1.4 μg/dL per 100 mg/kg), and above 300 mg/kg SL the BL exposure response was gradual (0.32 μg/dL per 100 mg/kg). The relationship between soil lead concentrations and blood lead levels in adults can be modelled using the USEPA Adult Lead Model (USEPA, n.d).

### Pregnant Individuals Exposure to Lead in Soil and Soil Derived Dust

Laidlaw et al. (2025) presented seven (7) studies where soil lead and indoor dust lead concentrations were correlated (P<0.05) and Laidlaw et al. (2014) described how lead in soil dust migrates into homes. Stanek et al. reported a statistically significant association between blood lead concentrations in pregnant individuals and lead levels measured from indoor surface wipe samples collected during pregnancy (Spearman r = 0.35, p < 0.0001). In an investigation of 24 households in Durham County, North Carolina, King et al. observed a positive correlation between residential soil lead concentrations and blood lead levels among pregnant individuals (r = 0.31, p = 0.0534).

### Lead Exposure in South Saharan Women

Women and children in SSA have been highly exposed to lead. Bede-Ojimadu et al. (2018) completed a systematic review of fifteen (15) studies that reported BLLs in women across SSA. The review reported mean blood lead concentrations among women varied widely, from 0.83 to 99 µg/dL, with an overall mean blood lead level was 24.73 µg/dL in women, and 26.24 µg/dL in pregnant women. In contrast, the central tendency of BLLs in women across North America, Europe, East Asia and Latin America, reported BLLs below approximately 1– 3 µg/dL, with most women in North America and Latin America having levels well under 1 µg/dL (see **Table S4**). Pregnancy represents a particularly vulnerable period as physiological changes can mobilize lead stored in bone, increasing maternal and fetal exposure (Gulson, 2003).

### Harmful Effects of Lead Exposure

Lead is a ubiquitous environmental toxicant with no level of exposure that is known to be without harmful effects (USEPA, 2024; WHO, 2026). The United States Environmental Protection Agency (US EPA) has evaluated the health consequences of lead exposure. Its Integrated Science Assessment for Lead (USEPA, 2024) describes causal relationships between lead exposure and adverse effects across multiple physiological systems. In children, neurological effects include reduced IQ and academic achievement, impaired learning, and attention deficits. In adults, lead exposure contributes to cognitive decline and poorer executive function, with potential implications for dementia-related outcomes. Cardiovascular effects include elevated blood pressure, hypertension, cardiovascular disease and cardiovascular mortality, while renal effects include reduced glomerular filtration rate and declining kidney function. Lead exposure is also causally associated with increased all-cause, non-accidental mortality. Biological responses, including oxidative stress and inflammation, provide mechanistic support for these effects. The assessment additionally identifies causal or likely causal relationships with reproductive and developmental outcomes, including impaired fetal growth, delayed puberty and reduced male reproductive function.

### Lead Exposure and Preeclampsia Studies

Three meta-analyses have reported large and statistically significant differences in maternal blood lead concentrations between women with preeclampsia and normotensive controls (**Table 3**). Poropat et al. (2018) reported a pooled Cohen’s d of 1.26 across 12 estimates involving 6,069 participants, while Zhong et al. (2022) reported a standardized mean difference (SMD) of 1.06 (95% CI 0.69–1.43) across 21 studies involving 12,531 participants. Notably, Zhong et al. also identified a statistically significant association in studies where mean blood lead concentrations in both groups were below 5 µg/dL (SMD 0.25; 95% CI 0.09– 0.40), suggesting that the association is not restricted to populations with markedly elevated lead exposure. Vigeh et al. (2025) subsequently reported a large pooled difference (SMD 2.19; 95% CI 0.91–3.48), although substantial between-study heterogeneity was observed. Collectively, these findings provide strong and consistent epidemiological evidence of an association between elevated maternal blood lead concentrations and preeclampsia, while heterogeneity, potential confounding, overlapping primary studies and limitations inherent in observational research preclude definitive causal inference.

**Table 3.** Statistics from three meta-analyses of maternal lead and preeclampsia.

| Meta-analysis | Studies and participants | Main pooled findings | Heterogeneity and qualifications |
| --- | --- | --- | --- |
| **Poropat et al. (2018)** | 11 eligible studies; 12 estimates; 6,069 participants | Cohen's d = 1.26; reported OR = 9.81 (interval 8.01–12.02); p = 0.005. Authors additionally reported a 1.6% increase in likelihood per 1 µg/dL increase in blood lead. | Moderate risk of bias in all studies. Removing one study produced a homogeneous analysis with stronger estimates. Numerical I<sup>2</sup> unavailable in accessible abstract. |
| **Zhong et al. (2022)** | 25 reviewed; 21 pooled; 12,531 participants: 1,533 cases and 10,998 controls | SMD = 1.06 (95% CI 0.69–1.43). In 11 studies with both groups' mean blood lead <5 µg/dL: SMD = 0.25 (95% CI 0.09–0.40; p = 0.002). | Overall I<sup>2</sup> = 96.4%; lower-exposure subgroup I<sup>2</sup> = 75%. Publication-bias tests: Begg's p = 0.001; Egger's p = 0.001. |
| **Vigeh et al. (2025)** | Nine studies: six on preeclampsia, three on gestational hypertension. Reported groups: 330 preeclampsia cases, 191 gestational-hypertension cases and 1,679 controls across all nine studies. | Mean blood lead: 18.4 µg/dL in preeclampsia versus 7.4 µg/dL in controls. Main text/discussion: SMD = 2.19 (95% CI 0.91–3.48; p <0.01). Abstract: SMD = 2.2 (95% CI 1.8–2.6). | Abstract heterogeneity by continent, design and sample size: 92.5%, 87.3% and 89.1%; main text reports different values. Egger's p <0.05. |

Notes: SMD = standardized mean difference; OR = odds ratio; CI = confidence interval. SMDs, including Cohen's d, are dimensionless differences in lead concentrations, not risk ratios or differences in µg/dL. The reviews contain overlapping primary studies.

### Association Between Soil Lead and Preeclampsia

In New Orleans, Louisiana Zahran et al., (2014) observed that mothers living in ZIP code areas where soil lead concentrations exceeded 333 mg/kg had a fourfold higher likelihood of eclampsia (OR = 4.00; 95% CI: 3.00–5.35) compared with those residing in neighborhoods with soil lead levels below 50 mg/kg. Laidlaw et al, (2025) discussed how lead in soil dust in urban areas could migrate into homes and potentially contribute to preeclampsia.

### Association Between Lead in Water and Preeclampsia

Lead levels in water have also been shown to be associated with the development of eclampsia (Troesken, 2006). Around 1900, Troesken (2006) observed that the mortality rate from eclampsia in counties with elevated lead levels in drinking water was 2.34 times higher than in counties with lower water-lead levels (95% CI: 1.54–3.14). This is the only known study that has documented the association between lead in water supplies and eclampsia or preeclampsia.

In this study, we estimate preeclampsia risk associated with observed maternal lead exposure levels in SSA using a comparative risk assessment approach. Our objective is not to predict individual clinical outcomes, but to quantify potential population-level risk in SSA associated with observed and modelled exposure scenarios relevant to the region.

## Methods

We conducted a population-level modelling study using a comparative risk assessment framework. Maternal BLLs were obtained from published biomonitoring studies conducted in sub-Saharan Africa (SSA) among pregnant women or women of reproductive age. To characterize extreme exposure contexts, additional scenarios were derived from reported soil lead concentrations in communities affected by mining, smelting, or informal lead–acid battery recycling and converted to estimated BLLs using established soil-to-blood lead modelling approaches.

The exposure–response relationship between blood lead and % preeclampsia risk was based on published meta-analytic evidence indicating a 1.6% increase in the odds of preeclampsia per 1 µg/dL increase in maternal blood lead level (Poropat et al., 2018). This relationship was implemented as a log-linear effect on the odds scale. Baseline preeclampsia risk (p0) was specified using plausible population-level prevalence estimates for sub-Saharan Africa and varied between 3% and 10% in sensitivity analyses. Figure S1 explains the interpretation of baseline prevalence, and Table SI presents the parameters and assumptions used in the comparative risk assessment of maternal lead exposure and preeclampsia

Predicted preeclampsia risk was estimated by converting baseline risk to odds, applying the lead-associated odds multiplier, and converting adjusted odds back to probability. Primary analyses capped BLLs at 100 µg/dL to avoid extrapolation beyond the epidemiologic evidence base. Uncapped analyses were conducted separately to illustrate risk saturation in extreme contamination scenarios.

Our analysis builds on the 15 studies summarized by Bede-Ojimadu et al. by incorporating additional pregnancy-specific biomonitoring data published since 2018 (see Table S2a).

### Overview of Comparative Risk Assessment Framework

This study applies a comparative risk assessment (CRA) framework consistent with Global Burden of Disease (GBD) methodology to estimate the contribution of maternal lead exposure to preeclampsia risk in sub-Saharan Africa. CRA quantifies the change in population risk associated with exposure relative to a specified baseline (counterfactual) risk, while holding other unmeasured determinants constant at their population-average levels.

### Exposure Definition

Exposure is defined as maternal blood lead level (BLL), expressed in micrograms per deciliter (µg/dL). BLLs were obtained either directly from pregnancy biomonitoring studies or estimated indirectly from soil lead concentrations using established soil-to-blood lead models for adult women. The theoretical minimum risk exposure level (TMREL) was assumed to be approximately 0–1 µg/dL, consistent with the absence of a known safe lead threshold.

### Exposure–Response Function

The exposure–response relationship was derived from meta-analytic evidence indicating a 1.6% increase in the odds of preeclampsia per 1 µg/dL increase in maternal BLL (Poropat et al., 2018). This relationship was implemented as a log-linear effect on the odds scale.

Relative odds as a function of BLL x were calculated as:

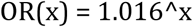

where OR(x) represents the odds of preeclampsia at blood lead level x relative to a reference exposure of 0 µg/dL.

### Baseline Risk (p0) Specification and Justification

Baseline preeclampsia risk (p0) represents the average probability of preeclampsia in the population in the absence of explicit modelling of lead exposure. Because individual-level clinical covariates (e.g., age, parity, body mass index, chronic hypertension) were unavailable, p0 serves as a population anchor that implicitly incorporates the distribution of these factors within the population.

Values of p0 between 3% and 10% were selected based on published estimates of preeclampsia prevalence and incidence in sub-Saharan Africa. Lower values (approximately 3–4%) reflect community-based and lower-risk populations, whereas higher values (6–10%) are consistent with facility-based and referral hospital settings common in the region.

Sensitivity analyses across this range were conducted to assess robustness to baseline uncertainty.

Conversion from Relative Odds to Absolute Risk Baseline odds were calculated from baseline risk p0 as:

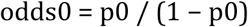

Lead-adjusted odds at exposure level x were then calculated as:

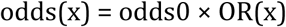

Predicted preeclampsia risk at exposure level x was obtained by converting odds back to probability:

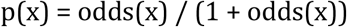

### Exposure Capping and Extreme Scenarios

To avoid extrapolation beyond the epidemiologic evidence base, primary analyses capped maternal blood lead levels at 100 µg/dL, corresponding to the upper range of most pregnancy biomonitoring studies. Uncapped analyses were conducted separately to illustrate risk saturation in extreme contamination scenarios (e.g., mining, smelting, informal battery recycling). Predicted risks approaching 100% in these scenarios are interpreted as indicators of model saturation rather than literal probabilities.

### Sensitivity analyses and population attributable fraction estimation

Sensitivity analyses were conducted to assess the influence of key modelling assumptions on predicted preeclampsia risk and population burden. First, to evaluate the impact of extrapolation beyond the epidemiologic evidence base, predicted risks were compared under capped and uncapped blood lead level (BLL) assumptions. In primary analyses, BLLs were capped at 100 µg/dL, corresponding to the upper range of pregnancy biomonitoring studies. Uncapped analyses extended the exposure–response function to extreme BLLs observed in severely contaminated environments to illustrate potential model saturation (**Figure 1**).

**Figure 1.**
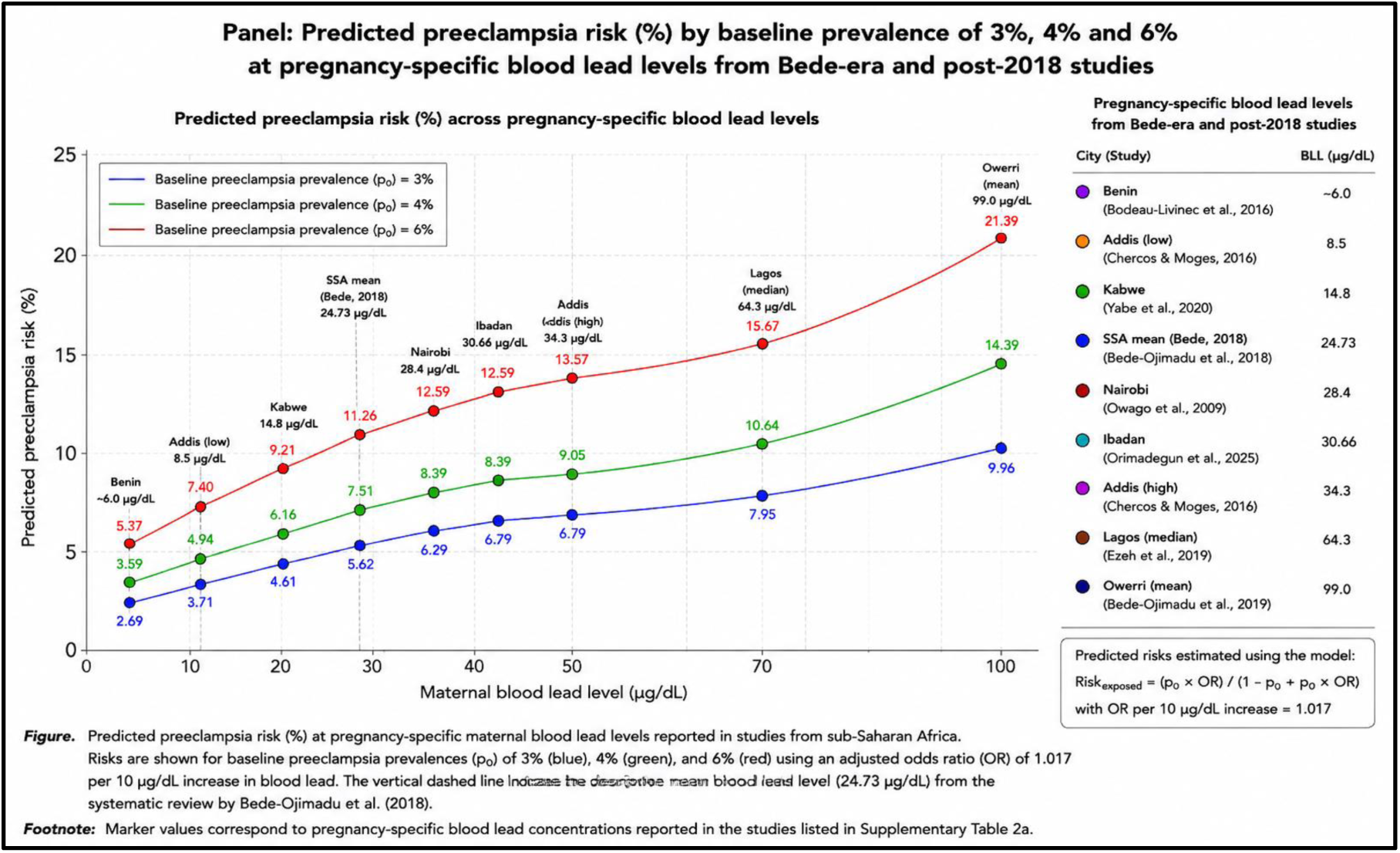
Predicted preeclampsia risk (%) at pregnancy-specific maternal blood lead levels reported in sub-Saharan Africa. Predicted preeclampsia risks are shown across pregnancy-specific maternal blood lead levels (BLLs) reported in Bede-era and post-2018 studies from sub-Saharan Africa. Risks are presented for baseline preeclampsia prevalence scenarios (p_0_) of 3% (blue), 4% (green), and 6% (red). Risks were estimated using a log-linear exposure–response model based on an adjusted odds ratio (OR) of 1.016 per 1 µg/dL increase in maternal blood lead level (equivalent to approximately OR 1.17 per 10 µg/dL increase). The vertical dashed line indicates the SSA mean maternal BLL (24.73 µg/dL) reported by Bede-Ojimadu et al. (2018). Markers represent pregnancy-specific maternal BLL concentrations reported in individual studies, including Bede-era and post-2018 biomonitoring studies (Supplementary Table S2a). **Footnote:** Marker values correspond to pregnancy-specific blood lead concentrations reported in the studies listed in Supplementary Table S2a. Predicted risks represent modelled estimates and assume that the observed association between maternal lead exposure and preeclampsia is causal.

Second, population attributable fractions (PAFs) for preeclampsia associated with maternal lead exposure were estimated across a range of baseline preeclampsia prevalence values representative of sub-Saharan Africa. PAFs were calculated within a comparative risk assessment framework using the modelled exposure distribution and the specified exposure–response relationship, allowing evaluation of how baseline prevalence assumptions influence absolute burden estimates.

### Adult soil lead to blood lead modelling

For communities with *severe* environmental lead contamination, maternal BLLs were estimated from soil lead concentrations using an adult soil-to-blood lead modelling approach consistent with the United States Environmental Protection Agency (US EPA) Adult Lead Methodology (ALM) (2003). The ALM estimates steady-state adult BLLs resulting from chronic environmental lead exposure and has been widely applied in contaminated-land risk assessment and environmental epidemiology.

Soil lead concentrations were converted to estimated maternal BLLs assuming chronic exposure through ingestion of contaminated soil and household dust, consistent with residential exposure scenarios (USEPA, 2003). The modelling assumed linear proportionality between external lead intake and blood lead concentration at environmentally relevant exposure levels, reflecting first-order biokinetic behavior in adults. Parameter values were selected to be conservative but plausible for women of childbearing age, including ingestion rates and bioavailability consistent with US EPA guidance.

The following assumptions were used in the soil to adult blood lead model: adult soil ingestion 50 mg/day; RBA 50%; adult blood lead slope factor (BKSF) 0.16 (µg/dL per µg/day). Thus Predicted BLL is approximately equal to Soil Pb × 0.004 (µg/dL per mg/kg).

Because pregnancy alters lead kinetics through mobilization of skeletal lead stores (Gulson et al., 2016), the estimated BLLs derived from soil concentrations should be interpreted as approximations of chronic exposure rather than precise pregnancy-specific predictions. Soil-derived scenarios were therefore used to illustrate potential exposure magnitudes in severely contaminated settings (e.g., mining, smelting, informal lead–acid battery recycling communities), rather than to represent typical population exposure.

To avoid extrapolation beyond the epidemiologic evidence base linking maternal BLL to preeclampsia, primary analyses capped modelled BLLs at values consistent with the upper range of pregnancy biomonitoring studies. Uncapped estimates were retained only for illustrative sensitivity analyses to demonstrate risk saturation under extreme contamination scenarios.

## Results

Maternal BLLs reported in sub-Saharan African biomonitoring studies ranged from near-background concentrations (<5 µg/dL) to more than 60 µg/dL in highly exposed urban and industrial settings. Across this range, increasing BLLs were associated with progressively higher relative odds of preeclampsia. Bede-Ojimadu et al. (2018) completed a systematic review of fifteen (15) studies that reported BLLs in women across SSA with a mean blood lead concentrations among women that varied widely, from 0.83 to 99 µg/dL. The overall mean blood lead level was 24.73 µg/dL. Using a mean BLL of 24.73 µg/dL translated into an absolute increases in predicted preeclampsia risk of approximately 4 to 8 percentage points at a p_o_ of 3% and 6%, respectively (see Figure 1A). Soil-derived exposure scenarios representing communities affected by severe environmental contamination yielded substantially higher modelled BLLs and correspondingly elevated predicted risks (see Table S3a).

Population biomonitoring programs in North America and Europe consistently report central tendency blood lead levels among women of reproductive from below 1 µg/dL to approximately 2.5 µg/dL, with most women having concentrations well below 1 µg/dL. In contrast, pregnancy-specific biomonitoring studies from SSA frequently report central tendency values seven-fold higher, with some facility-based and high-exposure settings reporting mean or median blood lead levels exceeding 10–20 µg/dL (Table S4). This disparity underscores the continued relevance of environmental lead exposure as a preventable contributor to maternal morbidity in the SSA region.

### Population Attributable Fraction

Population attributable fraction estimates varied with assumed baseline preeclampsia prevalence but preserved consistent exposure–response gradients across scenarios (Figure 2). Population attributable fractions were modestly sensitive to assumptions about baseline preeclampsia prevalence (p₀).

**Figure 2.**
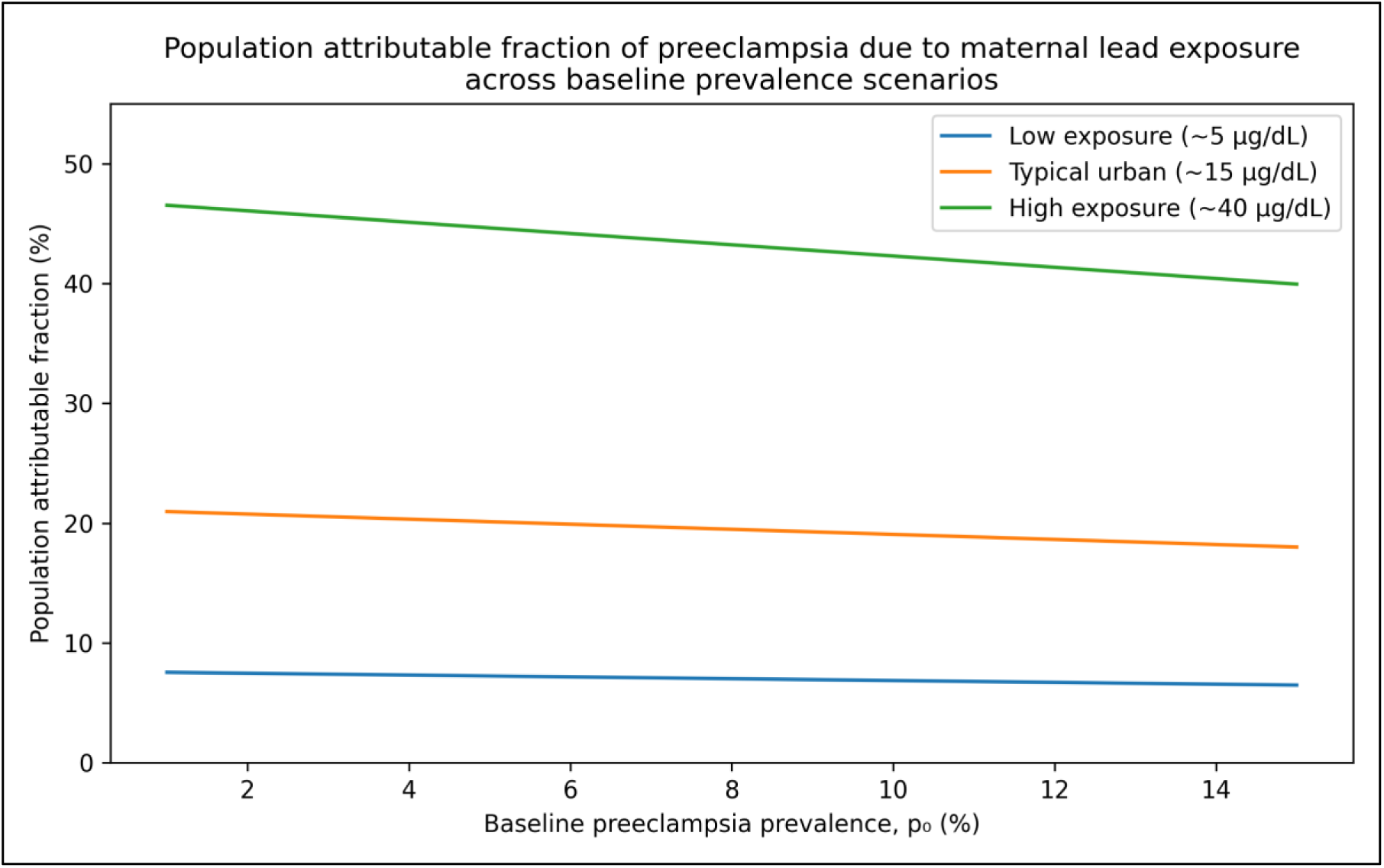
Population attributable fraction of preeclampsia due to maternal lead exposure across baseline prevalence scenarios. Population attributable fraction (PAF) of preeclampsia associated with maternal lead exposure plotted as a function of baseline preeclampsia prevalence (p₀). Estimates were derived using a comparative risk assessment framework assuming a log-linear increase in preeclampsia odds per 1 µg/dL increase in maternal blood lead level. Variation across baseline prevalence scenarios illustrates the dependence of absolute burden estimates on underlying population risk while preserving relative exposure–response gradients.

Higher baseline preeclampsia prevalence produced higher absolute predicted risks but lower population attributable fractions, because a greater proportion of the total disease burden was attributable to background risk. Conversely, lower baseline prevalence resulted in a larger proportional contribution of maternal lead exposure to the predicted preeclampsia burden. Thus, while absolute predicted preeclampsia risk increased with baseline prevalence, the estimated relative contribution of maternal lead exposure, expressed as the PAF, decreased as baseline prevalence increased. This reflects the mathematical dependence of risk-based PAF estimates on baseline risk. However, the relative contribution of maternal lead exposure remained substantial across all plausible prevalence scenarios, indicating that lead exposure is an important contributor to preeclampsia risk irrespective of baseline population risk.

Predicted preeclampsia risk represents the absolute probability of disease under a given exposure scenario, whereas the population attributable fraction quantifies the proportion of cases that can be attributed to lead exposure within that risk. These metrics provide complementary perspectives on individual risk and population-level impact.

## Discussion

This comparative risk assessment and previous work by Zahran et al (2014) in New Orleans, Louisiana (USA) and the review paper by Laidlaw et al., (2025), suggest that maternal lead exposure may contribute meaningfully to preeclampsia risk across sub-Saharan Africa. While the predicted increases in preeclampsia risk associated with typical biomonitoring-derived maternal blood lead levels (BLLs) are modest at the individual level, the widespread and persistent nature of exposure implies a potentially substantial population burden.

In New Orleans, Louisiana Zahran et al., (2014) observed that mothers living in ZIP code areas where soil lead concentrations exceeded 333 mg/kg had a fourfold higher likelihood of eclampsia (OR = 4.00; 95% CI: 3.00–5.35) compared with those residing in neighborhoods with soil lead levels below 50 mg/kg.

Importantly, elevated predicted risks were not limited to low level BLL scenarios. Maternal BLLs commonly reported in pregnancy biomonitoring studies across SSA are up to seven (7) fold higher than those observed in biomonitoring programs of women in western countries. The mean maternal blood lead level from fifteen (15) recent and separate studies in SSA was 26.24 µg/dL (Range: 0.83 – 99 µg/dL) (Bede-Ojimadu et al., 2018), while the mean or geometric mean BLL in of women the United States, Canada, Germany, South Korea, China, and Mexico were between 0.5 and 2.4 µg/dL. This highlights the SSA region as a global outlier for maternal lead exposure and underscoring the preventable nature of the associated preeclampsia burden.

In communities experiencing extreme environmental contamination such as Legacy mining and smelting sites and used lead acid battery sites, lead exposure may represent a dominant risk factor for preeclampsia, potentially overwhelming background maternal risk factors (see Tables 3a and 3b).

Thousands of abandoned or poorly regulated mine sites remain throughout the region (UNESCO, 2019), often in close proximity to residential areas that developed around historical operations. Soil lead concentrations at such sites frequently exceed international guideline values by several orders of magnitude, with well-documented examples including Kabwe (Zambia) (Nakayama, 2011) and mining-affected communities in Nigeria, South Africa, and the Central African Copperbelt. Despite their recognised public health importance, comprehensive population-relevant inventories linking contaminated mine sites to human exposure remain limited.

Informal used lead–acid battery (ULAB) processing represents another widespread source of extreme soil lead contamination. A global assessment estimated approximately 10,599– 29,241 informal ULAB processing sites across 90 countries in 2013, indicating a large and diffuse footprint (Ericson et al., 2016). However, region-specific inventories for sub-Saharan Africa remain poorly quantified, limiting systematic exposure assessment and prioritisation.

The concentration of exposure and attributable burden within a limited number of environments suggests that identifying and remediating severely lead-contaminated soils in densely populated settings may represent a particularly effective primary prevention strategy. Targeted environmental interventions in high-risk neighbourhoods have the potential to yield disproportionate maternal health benefits compared with diffuse exposure reduction approaches applied uniformly across entire cities or regions. Laidlaw et al. (2026) has evaluated various remedial solutions for diffuse lead in contaminated soil

The sensitivity analyses provide important context for interpreting the modelled risk estimates. Comparisons of capped and uncapped BLL scenarios demonstrate that extreme predicted risks arise primarily from extrapolation beyond the epidemiologic evidence base rather than from exposure levels typically observed among pregnant women. This supports the use of capped BLLs in primary analyses and reinforces the interpretation of uncapped results as illustrative of extreme contamination scenarios rather than literal individual-level risk predictions.

Analysis of population attributable fractions across baseline preeclampsia prevalence scenarios further highlights a key feature of comparative risk assessment: absolute burden estimates are dependent on underlying population risk, while relative exposure–response gradients are preserved. In settings with higher baseline prevalence of preeclampsia, such as referral hospitals or populations with limited access to antenatal care, the potential population burden attributable to lead exposure is correspondingly greater. Conversely, community-based settings with lower baseline prevalence yield smaller absolute attributable fractions despite similar relative risk relationships.

Together, these findings strengthen confidence in the qualitative conclusions of this study while underscoring the importance of transparent assumptions when translating exposure– response relationships into population-level burden estimates. By situating maternal blood lead exposure within its environmental context, this analysis highlights lead as a modifiable, preventable contributor to preeclampsia risk in sub-Saharan Africa and reinforces the need to integrate environmental source control with clinical maternal health strategies.

## Limitations

Limitations include reliance on published exposure–response estimates and the absence of individual-level covariates, which are subsumed within baseline risk assumptions. Nevertheless, the robustness of relative risk gradients across baseline scenarios supports the relevance of lead exposure as a population-level determinant of maternal health.

As with all comparative risk assessments, estimates presented here reflect population-level risk translation under explicit assumptions and should not be interpreted as individual-level causal predictions.

This study has several limitations that warrant careful consideration. First, the analysis is based on a comparative risk assessment framework and therefore provides population-level risk translation rather than individual-level clinical prediction. Individual maternal characteristics known to influence preeclampsia risk, including age, parity, body mass index, chronic hypertension, diabetes, and access to antenatal care, were not available at the individual level and are instead subsumed within the baseline preeclampsia risk parameter. While this approach is standard in comparative risk assessment and Global Burden of Disease–style analyses, it precludes adjustment for individual-level confounding and limits causal inference at the individual scale.

Second, the exposure–response relationship linking maternal blood lead level to preeclampsia risk was derived from published observational studies and meta-analyses (Poropat et al., 2018). Although these syntheses provide the best available quantitative evidence, residual confounding and exposure misclassification cannot be excluded. The assumed log-linear relationship on the odds scale may not hold uniformly across the full range of exposures considered, particularly at very high BLLs.

Third, data on maternal BLLs in SSA remain sparse and heterogeneous, with many studies based on small samples, urban settings, or specific exposure contexts. Soil-derived exposure scenarios used to represent communities affected by severe environmental contamination rely on modelling assumptions to translate soil lead concentrations into maternal BLLs. These scenarios are intended to be illustrative of extreme exposure conditions rather than representative of typical population exposure and should be interpreted with caution.

Fourth, to avoid extrapolation beyond the epidemiologic evidence base, primary analyses capped maternal BLLs at values consistent with the upper range of pregnancy biomonitoring studies. Uncapped analyses were conducted only to illustrate risk saturation under extreme contamination scenarios. Predicted risks approaching 100% in these contexts should not be interpreted as literal probabilities, but rather as indicators that exposure levels exceed those represented in available epidemiologic data and may overwhelm background maternal risk factors.

Finally, baseline preeclampsia prevalence varies widely across countries, healthcare settings, and study designs in SSA. Although sensitivity analyses across a broad range of plausible baseline risks demonstrated robustness of relative risk gradients, absolute predicted risks remain dependent on these baseline assumptions. Improved surveillance of both preeclampsia incidence and maternal lead exposure would strengthen future population-level risk assessments.

Despite these limitations, the transparent specification of assumptions, explicit sensitivity analyses, and consistency with established comparative risk assessment methods support the utility of this analysis for informing population-level prevention and policy prioritisation.

An additional limitation relates to the harmonization of soil and blood lead data. Soil lead and maternal BLL measurements were not collected contemporaneously or within the same individuals, and the alignment presented here is ecological and descriptive.

## Conclusions

In conclusion, this study suggests that maternal lead exposure may contribute meaningfully to preeclampsia risk in SSA, with modest effects at commonly observed exposure levels and potentially substantial effects in severely contaminated settings.

Given the preventable nature of lead exposure, addressing legacy urban soil contamination represents an under-recognised opportunity for primary prevention of maternal morbidity in the region.

### Recommendations

Prevention strategies for hypertensive disorders of pregnancy in SSA should extend beyond clinical management to include environmental surface soil lead exposure *isolation*. Priority should be given to identifying and isolating severely lead-contaminated soils in densely populated urban and peri-urban settings, particularly where women of childbearing age reside.

Low-cost, context-appropriate soil isolation and dust control strategies (Laidlaw et al, 2017) should be evaluated and implemented alongside routine maternal blood lead surveillance. Isolation of roadside soils may be a particular effective strategy (Laidlaw, 2014; Resongles et al., 2021). Integrating environmental source control with established clinical prevention measures, such as low-dose aspirin (Duley et al., 2017) and calcium supplementation (Hofmeyr et al. 2010), may yield synergistic reductions in preeclampsia risk.

Future research should prioritise integrated studies that concurrently measure soil lead, household dust, maternal blood lead, and pregnancy outcomes within the same communities to refine exposure–response estimates and inform targeted intervention strategies.

## Data Availability

All data produced in the present study are available upon reasonable request to the authors

## Acknowledgements

This study received no specific funding from public, commercial, or not-for-profit funding agencies.

## Ethics statements

Ethical approval was not required for this study.

## Patient and public involvement

Patients and the public were not involved in the design, conduct, reporting, or dissemination of this research.

## Declaration of generative AI and AI-assisted technologies in the manuscript preparation process

During the preparation of this work the author(s) used Chat GPT in order to perform calculations and prepare charts. After using this tool/service, the author(s) reviewed and edited the content as needed and take(s) full responsibility for the content of the published article.

## Supplementary Figure

**Figure S1.**
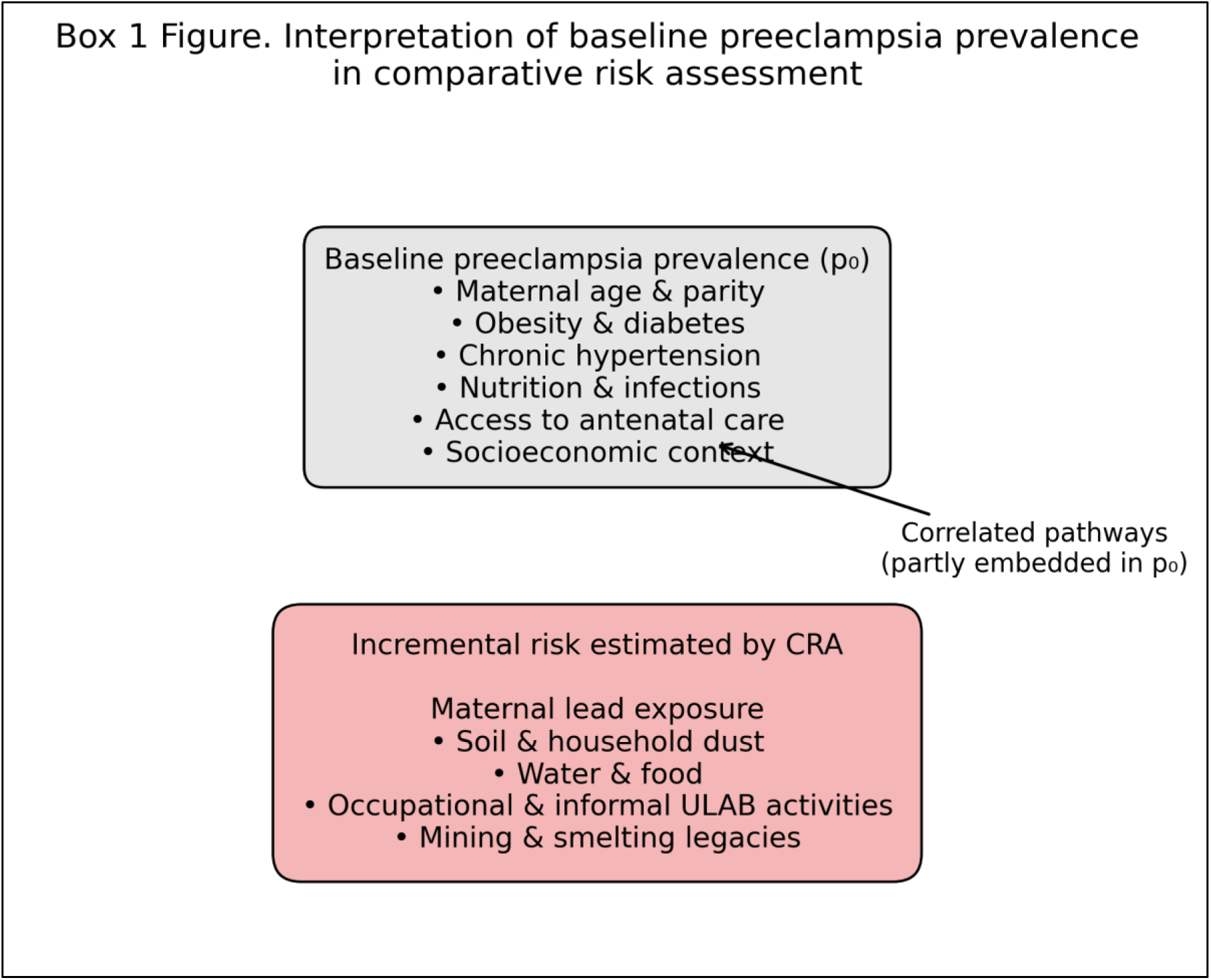
Conceptual framework for estimating incremental preeclampsia risk associated with maternal lead exposure. Baseline preeclampsia prevalence (p₀) represents underlying population risk determined by established maternal, clinical, nutritional, socioeconomic and health-system factors, including maternal age, parity, obesity, diabetes, hypertension and access to antenatal care (Duckett and Harrington, 2005; Abolos et al., 2013). Maternal lead exposure represents an incremental environmental risk component estimated using comparative risk assessment methods and published exposure–response evidence linking maternal blood lead levels with preeclampsia. Relevant exposure pathways include contaminated soil and household dust, drinking water and food, informal used lead-acid battery recycling, and mining/smelting-related contamination (Poropat et al., 2018; Laidlaw and Filippelli, 2008; Haefliger et al., 2009; Yabe et al., 2020).

## Supplementary Tables

**Table S1.** Parameters and assumptions used in the comparative risk assessment of maternal lead exposure and preeclampsia.

| Parameter | Value / assumption | Description | Source |
| --- | --- | --- | --- |
| **Exposure metric** | Maternal blood lead level (BLL, $\mu\text{g}/\text{dL}$) | Primary biomarker used to represent maternal lead exposure during pregnancy | Pregnancy biomonitoring studies from SSA |
| **Exposure-response relationship** | OR = 1.016 per 1 $\mu\text{g}/\text{dL}$ increase in maternal BLL | Estimated increase in odds of preeclampsia associated with increasing maternal BLL | Poropat et al. (2018) meta-analysis |
| **Equivalent exposure-response relationship** | OR = 1.17 per 10 $\mu\text{g}/\text{dL}$ increase in maternal BLL | Used for interpretation of exposure scenarios | Derived from Poropat et al. (2018) |
| **Baseline preeclampsia prevalence ($p_0$) scenarios** | 3%, 4%, 6% | Plausible background prevalence scenarios representing variation among populations | Abalos et al. (2013); SSA maternal health literature |
| **Low maternal lead exposure scenario** | 5 $\mu\text{g}/\text{dL}$ | Representative low exposure scenario | SSA biomonitoring distribution; contemporary international maternal BLLs |
| **Typical urban maternal lead exposure scenario** | 15 $\mu\text{g}/\text{dL}$ | Representative exposure in urban SSA settings | SSA pregnancy biomonitoring studies |
| **High maternal lead exposure scenario** | 40 $\mu\text{g}/\text{dL}$ | Representative high exposure/hotspot scenario | SSA pregnancy biomonitoring and contaminated community studies |
| **Extreme environmental exposure scenario** | >100 $\mu\text{g}/\text{dL}$ | Represents severely contaminated settings (e.g., mining or informal recycling communities) | SSA contaminated-site literature |
| **Risk model** | Log-linear odds model | Assumes a proportional increase in odds of preeclampsia with increasing maternal BLL | Poropat et al. (2018) |
| **Risk conversion** | $\text{Risk} = (p_0 \times \text{OR}) / (1 - p_0 + p_0 \times \text{OR})$ | Converts odds ratios into absolute predicted risks | Standard epidemiological risk conversion |
| **Population attributable fraction (PAF)** | $\text{PAF} = (\text{Risk}_{\text{exposed}} - p_0) / \text{Risk}_{\text{exposed}}$ | Estimates proportion of cases attributable to lead exposure under causal assumptions | Standard attributable risk methodology |
| **Biomonitoring dataset** | Pregnancy-specific SSA maternal BLL studies | Exposure distribution used for realistic exposure scenarios | Bede-Ojimadu et al. (2018) and subsequent studies |
| **Soil-derived scenarios** | Contaminated soil lead concentrations converted to maternal exposure scenarios | Used to evaluate severely contaminated environments not captured by biomonitoring studies | Exposure modelling assumptions described in Methods |
| **Causal assumption** | Association interpreted as potentially causal for PAF estimation | PAF estimates assume observed association between maternal lead exposure and preeclampsia is causal | Epidemiological convention |
| **Uncertainty assessment** | Sensitivity analyses varying baseline prevalence and exposure assumptions | Evaluates robustness of model outputs | Current study |

## Supplementary Tables (S2)

Bede-Ojimadu et al. (2018) identified 15 studies reporting blood lead levels among women of childbearing age in sub-Saharan Africa. Table S3a reconstructs these studies using information reported in the review and underlying primary publications. Table S3a extends this evidence base by incorporating pregnancy-specific biomonitoring studies published after 2018 or not captured by the original review, including large contemporary cohorts and studies conducted in known high-exposure settings.

**Table S2a.** Biomonitoring-derived maternal blood lead in sub-Saharan Africa reconstructed studies (Bede-Ojimadu et al., 2018) and Post 2018 studies.

| Evidence group | Country | City / setting | Survey year(s) | Population | Age (years) | n | BLL central tendency (µg/dL) | Dispersion / range (µg/dL) | Exposure source / context | Source URL |
| --- | --- | --- | --- | --- | --- | --- | --- | --- | --- | --- |
| Bede review | Zambia | Near lead mine | ≤1977 | Pregnant women living near a lead mine | NR | 122 | Mean 41.2 | SD 14.4 | Lead mining | Clark et al., 1977<br>https://doi.org/10.1136/pgmj.53.625.674 |
| Bede review | Zambia | Away from lead mine | ≤1977 | Pregnant women living away from a lead mine | NR | 31 | Mean 14.7 | SD 7.5 | Background/community comparison | Clark et al., 1977<br>https://doi.org/10.1136/pgmj.53.625.674 |
| Bede review | Nigeria | Ile-Ife | NR | Non-pregnant women of childbearing age occupationally exposed to lead | NR | 62 | Mean 6.81 | SD 2.61; range 2.46–15.09 | Occupational exposure; legacy petrol context | Ojo et al., 2007 |
| Bede review | Senegal | Dakar / Thiaroye-sur-Mer | 2007–2008 | Mothers of children who died of lead poisoning | 20–44 | 23 | Mean 55.3 | SD 19.8; range 32.5–98.8 | Informal used lead-acid battery recycling | Haefliger et al., 2009<br>https://doi.org/10.1289/ehp.0900696 |
| Bede review | Kenya | Nairobi | 1998 | Pregnant women attending two public prenatal clinics | 15–40 | 223 | Mean 28.4 | Range 0–295 | Urban/mixed; source not established | Odiambo et al., 1998<br>https://www.jofamericanscience.org/journals/am-sci/0503/06_0723.pdf |
| Bede review | South Africa | Rural area | 2005–2006 | Pregnant women | 14–41 | 96 | Median 2.09 | Range 0.74–5.03 | Rural environmental exposure | Rollin et al., 2009<br>https://doi.org/10.1039/B816236K |
| Bede review | South Africa | Urban area | 2005–2006 | Pregnant women | 14–41 | 96 | Median 3.29 | Range 1.63–8.15 | Urban/mixed | Rollin et al., 2009<br>https://doi.org/10.1039/B816236K |
| Bede review | South Africa | Industrial area | 2005–2006 | Pregnant women | 14–41 | 96 | Median 2.07 | Range 1.10–3.23 | Industrial | Rollin et al., 2009<br>https://doi.org/10.1039/B816236K |
| Bede review | South Africa | Atlantic Ocean area | 2005–2006 | Pregnant women | 14–41 | 96 | Median 2.37 | Range 1.06–3.89 | Coastal/background | Rollin et al., 2009<br>https://doi.org/10.1039/B816236K |
| Bede review | South Africa | Mining area | 2005–2006 | Pregnant women | 14–41 | 96 | Median 2.64 | Range 0.61–16.15 | Mining | Rollin et al., 2009<br>https://doi.org/10.1039/B816236K |
| Bede review | South Africa | Indian Ocean area | 2005–2006 | Pregnant women | 14–41 | 96 | Median 2.19 | Range 0.88–2.94 | Coastal/background | Rollin et al., 2009<br>https://doi.org/10.1039/B816236K |
| Bede review | South Africa | Inland area | 2005–2006 | Pregnant women | 14–41 | 96 | Median 1.15 | Range 1.63–4.94 (as reported in review) | Inland/background | Rollin et al., 2009<br>https://doi.org/10.1039/B816236K |
| Bede review | Nigeria | Lagos | 2006–2008 | Pregnant women | 17–49 | 317 | Mean 59.5 | SE 2.1 | Urban/mixed | Adkunle et al., 2009<br>https://doi.org/10.1007/s10661-009-1247-4 |
| Bede review | Nigeria | Lagos | 2006–2008 | Non-pregnant women | 17–49 | 317 | Mean 27.7 | SE 1.1 | Urban/mixed | Adkunle et al., 2009<br>https://doi.org/10.1007/s10661-009-1247-4 |
| Bede review | Nigeria | Edo | 2006–2008 | Women with preeclampsia | NR | 59 | Mean 60.2 | SD 12.8 | Urban/mixed; source not reported | Ikechukwu et al., 2012<br>https://doi.org/10.1080/19338244.2011.619212 |
| Bede review | Nigeria | Edo | 2006–2008 | Normal pregnant women | NR | 150 | Mean 26.3 | SD 8.0 | Urban/mixed; source not reported | Ikechukwu et al., 2012<br>https://doi.org/10.1080/19338244.2011.619212 |
| Bede review | Nigeria | Edo | 2006–2008 | Non-pregnant women | NR | 122 | Mean 13.1 | SD 6.4 | Urban/mixed; source not reported | Ikechukwu et al., 2012<br>https://doi.org/10.1080/19338244.2011.619212 |
| Bede review | Nigeria | Owerri | 2011 | Pregnant women | NR | 99 | Mean 99.0 | SD 123; range 2–448 | Rural/urban comparison; mixed environmental exposure | Njoku and Orisakwe, 2012<br>https://doi.org/10.1136/oemed-2012-100947 |
| Bede review | Nigeria | Abakaliki | 2007–2008 | Pregnant women, gestational age ≤25 weeks | 15–40 | 349 | Mean 36.4 | SD 18.5; range 2.7–73.8 | Urban/mixed; source not reported | Ugwula et al., 2012<br>https://doi.org/10.1007/s10661-012-2828-1 |
| Bede review | Botswana | Central District | 2009–2010 | First-trimester pregnant women | 18–44 | 137 | Mean 1.96 | SE 0.14 | Community; source not reported | Mgowe et al., 2013 |
| Bede review | Botswana | Central District | 2009–2010 | Second-trimester pregnant women | 18–44 | 126 | Mean 2.49 | SE 0.17 | Community; source not reported | Mgowe et al., 2013 |
| Bede review | Botswana | Central District | 2009–2010 | Third-trimester pregnant women | 18–44 | 106 | Mean 2.66 | SE 0.19 | Community; source not reported | Mgowe et al., 2013 |
| Bede review | South Africa | Johannesburg | 2010 | All pregnant women | 18–46 | 306 | Geometric mean 0.83 | NR | Urban/mixed | Mathee et al., 2014<br>http://dx.doi.org/10.7196/samj.7466 |
| Bede review | South Africa | Johannesburg | 2010 | Non-geophagic pregnant women | 18–46 | 247 | Geometric mean 1.44 | Range 1.0–9.9 | Urban/mixed | Mathee et al., 2014<br>http://dx.doi.org/10.7196/samj.7466 |
| Bede review | South Africa | Johannesburg | 2010 | Geophagic pregnant women | 18–46 | 60 | Geometric mean 2.06 | Range 1.0–8.6 | Geophagia / direct soil ingestion | Mathee et al., 2014<br>http://dx.doi.org/10.7196/samj.7466 |
| Bede review | Nigeria | Nnewi | 2010–2011 | Women at delivery | 18–40 | 119 | Mean 6.19 | SD 2.77; range 2.17–15.25 | Urban/mixed; source not reported | Obi et al., 2014<br>https://doi.org/10.7888/juoeh.36.159 |
| Bede review | Ethiopia | Addis Ababa–Adama highway | 2011 | Women living along a highway | Mean 27.6 ± 7.2 | 40 | Mean 34.32 | SD 6.69 | Traffic / highway exposure | Chercos and Moges, 2016<br>https://doi.org/10.1155/2016/7569157 |
| Bede review | Ethiopia | ≈10 km from Addis Ababa–Adama highway | 2011 | Women living away from highway | Mean 26.3 ± 6.3 | 36 | Mean 8.47 | SD 3.01 | Lower-traffic comparison | Chercos and Moges, 2016<br>https://doi.org/10.1155/2016/7569157 |
| Bede review | Benin | Cotonou | 2011–2013 | Mothers of children aged 1–2 years with elevated BLL | NR | 227 | Mean 5.14 | SD 2.23; range 2.28–20.20 | Mixed household/environmental sources | Bodeau-Livinec et al., 2016<br>https://doi.org/10.3390/ijerph13030316 |
| Post-2018 update | Nigeria | Lagos | 2018 | Mothers at delivery | NR | 400 | Median 64.3 | Range 0–332 | Urban/mixed | Ladele et al., 2019<br>https://doi.org/10.1371/journal.pone.0211535 |
| Post-2018 update | Zambia | Kabwe | 2017 | Mothers / adult women in contaminated households | NR | NR | Mean 14.8 | NR | Lead–zinc mining and smelting legacy | Yabe et al., 2020<br>https://doi.org/10.1016/j.chemosphere.2019.125412 |
| Post-2018 update | Tanzania | Artisanal and small-scale gold-mining communities | 2015–2017 | Pregnant women in mining communities | NR | 883 | Median 2.722 | Reported in original study | Artisanal gold mining and geophagia | Nyanza et al., 2021<br>https://doi.org/10.1016/j.envint.2020.106104 |
| Post-2018 update | Tanzania | Non-mining comparison communities | 2015–2017 | Pregnant women | NR | 173 | Median 2.032 | Reported in original study | Community comparison | Nyanza et al., 2021<br>https://doi.org/10.1016/j.envint.2020.106104 |
| Post-2018 update | Nigeria | Ibadan | 2023 | Pregnant women | 18–49 | 78 | Mean 30.66 | SD 25.8 | Urban/mixed; environmental source not measured | Orimadegun et al., 2026<br>https://doi.org/10.1007/s12011-025-04703-0 |

**Table S3a.**
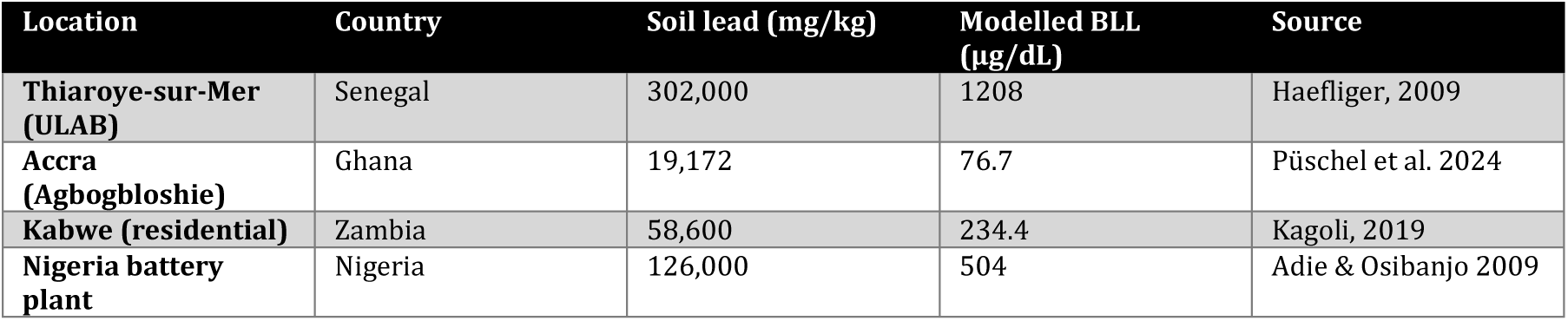
Severe soil-lead contamination derived exposure scenarios and modelled maternal blood lead levels.

| Location | Country | Soil lead (mg/kg) | Modelled BLL (µg/dL) | Source |
| --- | --- | --- | --- | --- |
| Thiaroye-sur-Mer (ULAB) | Senegal | 302,000 | 1208 | Haefliger, 2009 |
| Accra (Agbogbloshie) | Ghana | 19,172 | 76.7 | Püschel et al. 2024 |
| Kabwe (residential) | Zambia | 58,600 | 234.4 | Kagoli, 2019 |
| Nigeria battery plant | Nigeria | 126,000 | 504 | Adie & Osibanjo 2009 |

**Table S3b.** Relative odds and predicted preeclampsia risk across baseline scenarios.

| Maternal BLL (µg/dL) | Relative odds (1.016^BLL) | PE risk (%) p0=3% | PE risk (%) p0=4% | PE risk (%) p0=6% | PE risk (%) p0=10% |
| --- | --- | --- | --- | --- | --- |
| 5 | 1.08 | 3.2 | 4.3 | 6.4 | 10.6 |
| 10 | 1.17 | 3.4 | 4.6 | 6.9 | 11.4 |
| 20 | 1.37 | 4.1 | 5.3 | 7.9 | 13.0 |
| 40 | 1.89 | 5.5 | 7.3 | 10.8 | 17.3 |
| 60 | 2.59 | 7.4 | 9.8 | 14.2 | 22.4 |
| 80 | 3.56 | 9.9 | 12.9 | 18.5 | 28.3 |
| 100 | 4.89 | 13.1 | 16.9 | 23.8 | 35.2 |

**Table S4.**
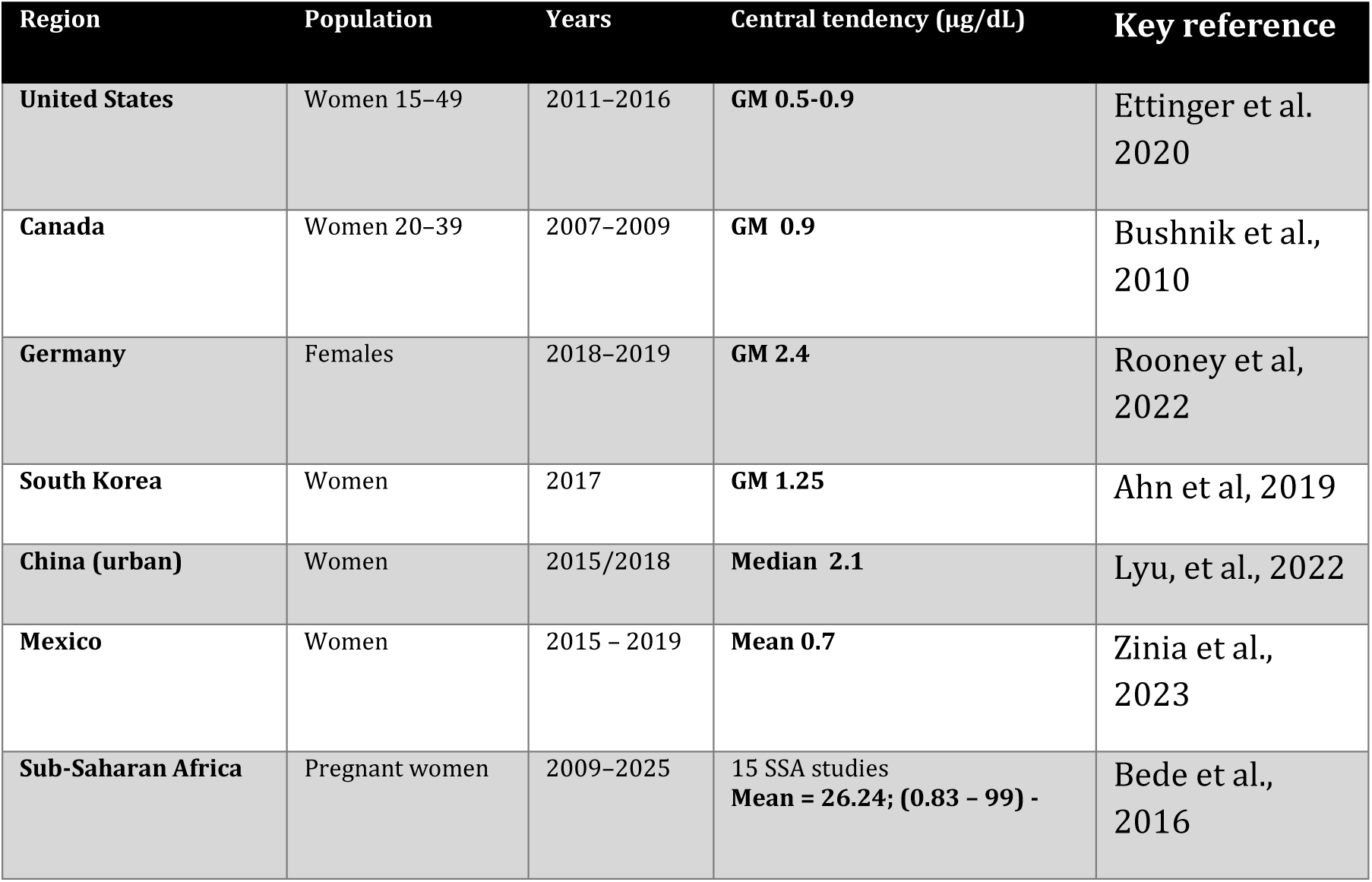
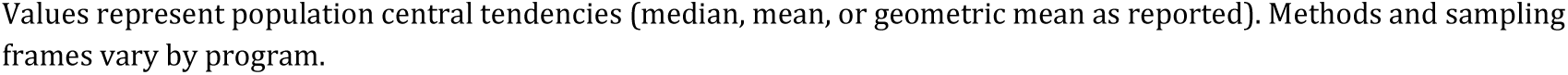
Central tendency of blood lead levels among women of reproductive age in selected world regions (≈2007–2025)

| Region | Population | Years | Central tendency (µg/dL) | Key reference |
| --- | --- | --- | --- | --- |
| United States | Women 15–49 | 2011–2016 | GM 0.5–0.9 | Ettinger et al. 2020 |
| Canada | Women 20–39 | 2007–2009 | GM 0.9 | Bushnik et al., 2010 |
| Germany | Females | 2018–2019 | GM 2.4 | Rooney et al, 2022 |
| South Korea | Women | 2017 | GM 1.25 | Ahn et al, 2019 |
| China (urban) | Women | 2015/2018 | Median 2.1 | Lyu, et al., 2022 |
| Mexico | Women | 2015 – 2019 | Mean 0.7 | Zinia et al., 2023 |
| Sub-Saharan Africa | Pregnant women | 2009–2025 | 15 SSA studies<br>Mean = 26.24; (0.83 – 99) - | Bede et al., 2016 |

Values represent population central tendencies (median, mean, or geometric mean as reported). Methods and sampling frames vary by program.

